# Longitudinal multi-omics profiling of tofacitinib response identifies a pre-existing Janus kinase-tractable inflammatory state in ulcerative colitis

**DOI:** 10.64898/2026.10.07.26364964

**Authors:** Ivan C.N. Fung, Djuna C. de Jong, Ke Liu, Andrew Y.F. Li Yim, Sara van Gennep, Rishand K. Ramkisoen, Esmé Clasquin, Jitteke de Jong, Krisztina B. Gecse, Mark Löwenberg, Aart Mookhoek, Wouter J. de Jonge, Geert R. D’Haens

## Abstract

**Background & Objective:** Tofacitinib, a Janus kinase (JAK) inhibitor, is approved for ulcerative colitis (UC). We investigated baseline and longitudinal transcriptomics and proteomics in intestinal biopsies, peripheral blood, and serum from UC patients starting tofacitinib to characterize treatment-associated effects and identify which molecular compartment harbors the most predictive response-associated biomarkers.

**Methods:** In this prospective, open-label study, patients with moderately to severely active UC started tofacitinib 10 mg BID for eight weeks. Clinical, biochemical, endoscopic, and histological data were collected at baseline and week 8. The primary outcome, histo-endoscopic mucosal improvement (HEMI), was defined as endoscopic Mayo subscore ≤1 with histologic remission (Robarts Histopathology Index ≤3, no mucosal neutrophils). Mucosal biopsies and peripheral blood were collected at baseline and week 8 for bulk RNA-sequencing (RNA-seq); biopsies and serum underwent proteomic profiling using Olink Explore 384 (Cardiometabolic, Inflammation, Neurology, Oncology panels).

**Results:** In total, 127 paired multi-omic samples were collected longitudinally from 40 UC patients: 15 (38%) achieved HEMI at week 8 (responders), 13 (32.5%) were late responders, and 12 (30%) were non-responders. Pretreatment mucosal biopsies from future responders showed elevated interferon, IL-6-family, IL-2/common-γ-chain, and T-cell-receptor signaling gene expression, which declined on treatment only in responders. A responder-specific JAK-STAT-dependent mechanism spanned interferon, cytokine-receptor, T-cell-receptor, and NF-κB signaling, with concordant gene- and protein-level reductions. Compared with a public infliximab dataset, JAK-STAT/interferon downregulation was shared and response-independent, while non-canonical NF-κB downregulation was tofacitinib-specific. In blood, shared pathway labels reflected distinct genes from mucosal effectors.

**Conclusion:** Response to tofacitinib depends on a JAK-tractable mucosal pathway absent in non-responders, suggesting a pretreatment biomarker for predicting treatment response.

*What is already known on this topic:* Prior studies have individually interrogated transcriptomic and/or epigenomic signatures of tofacitinib response as a single modality.

*What this study adds:* Our study provides simultaneous profiling of both the transcriptome and proteome across paired mucosal and systemic compartments in patients, comparing responders with non-responders at baseline and over time. Our data support a model where tofacitinib response in UC depends on a JAK-dependent pro-inflammatory phenotype that the drug can dismantle, in contrast to non-responders representing distinct, JAK-independent pathways.

*How this study might affect research, practice or policy:* This study provides a foundation for treatment response studies in UC towards the development and validation of predictive biomarkers in colonic mucosa and blood, which could ultimately result in more personalized treatment strategies.

## INTRODUCTION

Tofacitinib is an orally administered small molecule inhibitor of the Janus kinase (JAK) family that targets JAK1 and JAK3 with residual activity against JAK2 and TYK2[1]. By blocking the phosphorylation and activation of signal transducer and activator of transcription (STAT) proteins, tofacitinib impairs downstream signaling pathways involved in the transcription of pro-inflammatory cytokines, such as interleukin (IL)-2, IL-6, IL-12, and interferon (IFN)-γ[2], thereby interfering in key immune processes that drive chronic inflammation in immune-mediated diseases[3]. Tofacitinib has demonstrated safety and efficacy in inducing and maintaining remission in patients with moderate-to-severe UC [4, 5, 6], with rapid symptom improvement observed as early as three days after treatment initiation[7].

Despite its broad immunosuppressive effects, a recent meta-analysis indicated that clinical response rates in UC patients starting treatment with tofacitinib were 58% and 51% at week 8 and week 52, respectively[8], underscoring the need for predictive biomarkers and a deeper molecular and immunological understanding of treatment efficacy[9, 10, 11]. Using exposure-response modelling across the pivotal OCTAVE registration trial, Mukherjee *et al*.[11] demonstrated that prior anti-TNF failure was a significant clinical covariate negatively associated with therapeutic efficacy, with patients naïve to anti-TNF therapy showing substantially higher remission rates at both induction and maintenance time points. At the molecular level, we previously identified a panel of 53 differentially methylated positions (DMPs) in peripheral blood obtained pretreatment, that were associated with future endoscopic and clinical response, achieving an area under the receiver operating characteristic of 0.74[9]. More recently, Melón-Ardanaz *et al*. 2025[10] applied single-cell RNA sequencing (scRNA-seq) on intestinal biopsies collected before treatment initiation with tofacitinib, demonstrating that enhanced baseline JAK-STAT pathway activity in T cells, B cells, mast cells, fibroblasts, and endothelial cells identified future responders, while elevated baseline NF-κB activity in T cells, macrophages, and epithelial cells characterized non-responders[10]. Importantly, therapeutic resistance to tofacitinib was mechanistically linked to hyperactivation of myeloid cells, with IL-10-dependent macrophages identified as a key cellular subset contributing to treatment failure. Collectively, these findings highlight the potential of both molecular and cellular pretreatment signatures to inform patient selection and advance precision medicine approaches in UC.

While prior studies have individually interrogated transcriptomic and/or epigenomic signatures of tofacitinib response, the simultaneous profiling of both the transcriptome and proteome across paired mucosal and systemic compartments in the same patients remains unexplored. By integrating bulk RNA-sequencing (RNA-seq) and Olink proteomics from mucosal biopsies and peripheral blood over time, the present study provides a comprehensive, cross-compartment view of the molecular programs that distinguish responders from non-responders, both pretreatment and during treatment.

## METHODS

### Study design and population

We performed a single-center, prospective, open-label study enrolling adult patients with moderate-to-severe active UC, defined by a total Mayo score ≥ 6 and an endoscopic Mayo (eMayo) subscore ≥2. Patients were treated with tofacitinib 10mg BID for a minimum of eight weeks, with enrolment occurring between December 2017 and June 2020. Eligible patients were between 18 and 65 years of age and had a confirmed diagnosis of UC at least four months prior to inclusion. Prior treatment with thiopurines, corticosteroids, and biological agents were allowed. Concomitant use of oral corticosteroids (prednisone, budesonide) and oral aminosalicylates at stable doses for at least 2 and 4 weeks, respectively, was allowed. Patients were excluded if they received concomitant topical therapies (e.g. enemas and suppositories), immunomodulatory agents (e.g. thiopurines or methotrexate) or biologicals.

### Response characterization

This work represents a substudy of the TOFA-histology trial, where patients achieving histo-endoscopic mucosal improvement (HEMI) at week 8 were classified as responders (R)[12]. HEMI was defined as an eMayo subscore ≤ 1 in combination with histological remission as determined by a Robarts Histopathology Index (RHI) ≤ 3 in the absence of mucosal neutrophils. Patients that did not achieve HEMI at week 8 were classified as non-responders (NR). While a subset of the non-responders did not achieve HEMI at week 8, they showed clinical improvement and accordingly continued tofacitinib for at least one year demonstrating endoscopic improvement at subsequent assessment during routine care. We therefore classified this subset of NR as late responders (LR). Other endpoints can be found in van Gennep *et al*.^[12]^.

### Patient sampling

As detailed in Gennep *et al*.[12], the study protocol comprised six outpatient visits over a 12-week follow-up period. Following completion, patients resumed routine care, with an additional follow-up visit at approximately week 52. Sigmoidoscopy was performed at screening and at week 8. Patients demonstrating clinical benefit at week 8 underwent a further endoscopy at week 52 as part of standard care. At screening, five colonic biopsies (two for hematoxylin and eosin histology, one for RNA-seq, and two for Olink) were taken from the most inflamed mucosa, 15-30 cm from the anal verge. Another three colonic biopsies were acquired from normal appearing mucosa (two for histology and one for RNA-seq) higher up in the colon. Follow-up biopsies were collected at week 8 from the same anatomical location, defined by the distance from the anal verge, regardless of the endoscopic appearance at that point in time. Peripheral blood and serum were collected at screening and week 8. Whole blood was collected in PAXgene blood tubes and frozen at −20°C for 24 hours before storage at −80°C until analysis. Serum was collected using serum separation tubes, centrifuged after clotting, and stored at −80°C until analysis.

The total Mayo score was assessed at baseline and week 8, with partial Mayo scores recorded at weeks 2 and 4. Adverse events, laboratory results, concomitant medication use, and treatment adherence were monitored at each visit and through patient diaries. A gastroenterologist blinded to patient history assessed endoscopic disease severity on recorded videos using the eMayo subscore (0-3 point scale) and ulcerative colitis endoscopic index of severity (UCEIS; 0-8 point scale) for each visualized colonic segment. Histological inflammation was quantified by an IBD pathologist blinded to patient history using the RHI (0-33 point scale) and the converted Geboes score (GS; 0-22 point scale). Mean RHI and GS values were calculated from biopsies collected from the same anatomical locations.

### Sample processing

Colonic biopsies designated for histological analysis were fixated in 10% formalin and processed for hematoxylin and eosin (H&E) staining. Biopsies for RNA analysis were transferred to RNAlater and stored at 4°C for 24 hours prior to long-term storage. Protein samples were stored in 10% DMSO and slow-frozen using a controlled-rate freezing container (Mr. Frosty). Colonic biopsies and blood samples were stored at −80°C until analysis.

### Transcriptomic, proteomic, and computational analyses

Bulk RNA-seq was performed on RNA extracted from mucosal biopsies and peripheral whole blood collected at baseline and week 8, with differential expression and pathway enrichment analyses used to characterize treatment- and response-associated transcriptional changes. Protein profiling of matched mucosal biopsies and serum was performed using the Olink explore 384 cardiometabolic, inflammation, neurology, and oncology panels. To contextualize tofacitinib-associated transcriptional changes against another mechanism of action and similar dataset comparison, publicly available processed infliximab bulk transcriptomic microarray data from GSE23597[13] and tofacitinib single-cell RNA-sequencing (scRNA-seq) from GSE253006[10] were acquired for validation and specificity purposes. Supervised machine learning was applied across the transcriptomic and proteomic datasets to identify the molecular compartment most predictive of treatment response. Full details of RNA-seq and Olink sample processing, sequencing and proteomic platforms, bioinformatic pipelines, differential expression and pathway analysis methods, public dataset reprocessing, and machine learning model development are provided in **Supplementary Information**.

### Statistical analysis of clinical parameters

Continuous variables were expressed as mean ± standard deviation (SD) or median and interquartile range (IQR), according to distribution. Categorical variables were expressed as numbers with percentages. Normally distributed continuous data were compared by using the independent-sample t-test. If continuous data were not normally distributed, the Mann-Whitney U test was applied. Categorical variables were compared using the Pearson chi-square test. SPSS 28.0 (IBM Corporation) was used to perform the statistical analyses, with a predefined two-sided significance limit of *p* < 0.05. Detailed analyses of predictors for tofacitinib response and immunohistochemical assessments are presented in our separate manuscript[12].

### Ethical statement

This study was approved by the Medical Ethics Committee of the Academic Medical Center (METC2017_120) and registered in the Dutch Trial Register (https://www.onderzoekmetmensen.nl/) under trial number NL-OMON47601. All patients provided written informed consent prior to inclusion. All authors had access to the study data and reviewed and approved the final manuscript.

### Data availability statement

Processed RNA-sequencing counts as well as the OLINK NPX values can be found on Zenodo DOI: 10.5281/zenodo.23210920. The full Snakemake pipeline can be found on Github at https://github.com/AmsterdamUMC/ELNID001000000000496617.

## RESULTS

### Patient population and clinical outcome

In total, 40 patients were included, of which the majority (65%) presented with severe endoscopic disease (eMayo subscore of 3), involving at least the left colon (E2: 53%, E3: 43%) (**Table 1**). Paired inflamed mucosal biopsies, peripheral blood (PB) and serum were collected and analyzed (**Figure 1A**). Prior treatment included anti-TNF (78%), vedolizumab (55%), or both (43%). Fifteen patients (37.5%) achieved HEMI at week 8, which we termed responders (R). Twenty five patients (62.5%) were non-responders (NR). Out of these patients, thirteen (32.5%) achieved long-term clinical remission (**Figure 1B**), which we termed late responders (LR). Comparison of baseline characteristics revealed no significant differences between responders and non-responders, except for prednisone use and CRP levels, which were significantly higher in responders and non-responders, respectively (**Table 1**).

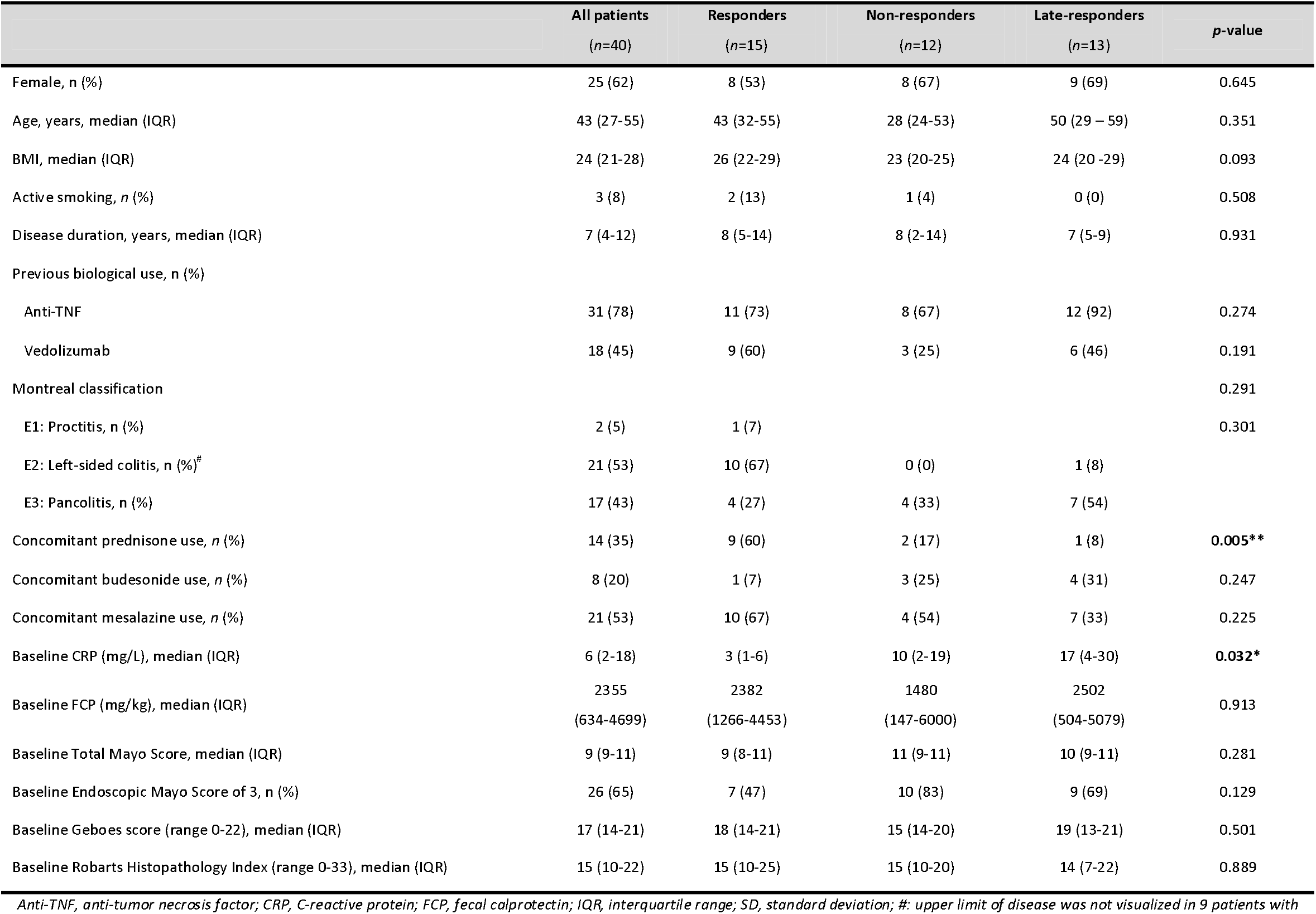
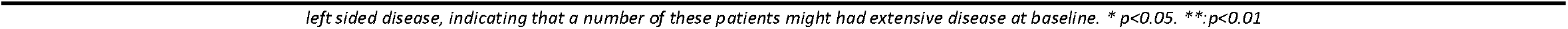

|  | All patients<br>(n=40) | Responders<br>(n=15) | Non-responders<br>(n=12) | Late-responders<br>(n=13) | p-value |
| --- | --- | --- | --- | --- | --- |
| Female, n (%) | 25 (62) | 8 (53) | 8 (67) | 9 (69) | 0.645 |
| Age, years, median (IQR) | 43 (27-55) | 43 (32-55) | 28 (24-53) | 50 (29 – 59) | 0.351 |
| BMI, median (IQR) | 24 (21-28) | 26 (22-29) | 23 (20-25) | 24 (20 -29) | 0.093 |
| Active smoking, n (%) | 3 (8) | 2 (13) | 1 (4) | 0 (0) | 0.508 |
| Disease duration, years, median (IQR) | 7 (4-12) | 8 (5-14) | 8 (2-14) | 7 (5-9) | 0.931 |
| Previous biological use, n (%) |  |  |  |  |  |
| Anti-TNF | 31 (78) | 11 (73) | 8 (67) | 12 (92) | 0.274 |
| Vedolizumab | 18 (45) | 9 (60) | 3 (25) | 6 (46) | 0.191 |
| Montreal classification |  |  |  |  | 0.291 |
| E1: Proctitis, n (%) | 2 (5) | 1 (7) |  |  | 0.301 |
| E2: Left-sided colitis, n (%)<sup>#</sup> | 21 (53) | 10 (67) | 0 (0) | 1 (8) |  |
| E3: Pancolitis, n (%) | 17 (43) | 4 (27) | 4 (33) | 7 (54) |  |
| Concomitant prednisone use, n (%) | 14 (35) | 9 (60) | 2 (17) | 1 (8) | **0.005\*\*** |
| Concomitant budesonide use, n (%) | 8 (20) | 1 (7) | 3 (25) | 4 (31) | 0.247 |
| Concomitant mesalazine use, n (%) | 21 (53) | 10 (67) | 4 (54) | 7 (33) | 0.225 |
| Baseline CRP (mg/L), median (IQR) | 6 (2-18) | 3 (1-6) | 10 (2-19) | 17 (4-30) | **0.032\*** |
| Baseline FCP (mg/kg), median (IQR) | 2355<br>(634-4699) | 2382<br>(1266-4453) | 1480<br>(147-6000) | 2502<br>(504-5079) | 0.913 |
| Baseline Total Mayo Score, median (IQR) | 9 (9-11) | 9 (8-11) | 11 (9-11) | 10 (9-11) | 0.281 |
| Baseline Endoscopic Mayo Score of 3, n (%) | 26 (65) | 7 (47) | 10 (83) | 9 (69) | 0.129 |
| Baseline Geboes score (range 0-22), median (IQR) | 17 (14-21) | 18 (14-21) | 15 (14-20) | 19 (13-21) | 0.501 |
| Baseline Roberts Histopathology Index (range 0-33), median (IQR) | 15 (10-22) | 15 (10-25) | 15 (10-20) | 14 (7-22) | 0.889 |

Anti-TNF, anti-tumor necrosis factor; CRP, C-reactive protein; FCP, fecal calprotectin; IQR, interquartile range; SD, standard deviation; #: upper limit of disease was not visualized in 9 patients with
*left sided disease, indicating that a number of these patients might had extensive disease at baseline. \* $p < 0.05$ . \*\*: $p < 0.01$*

**Figure 1.**
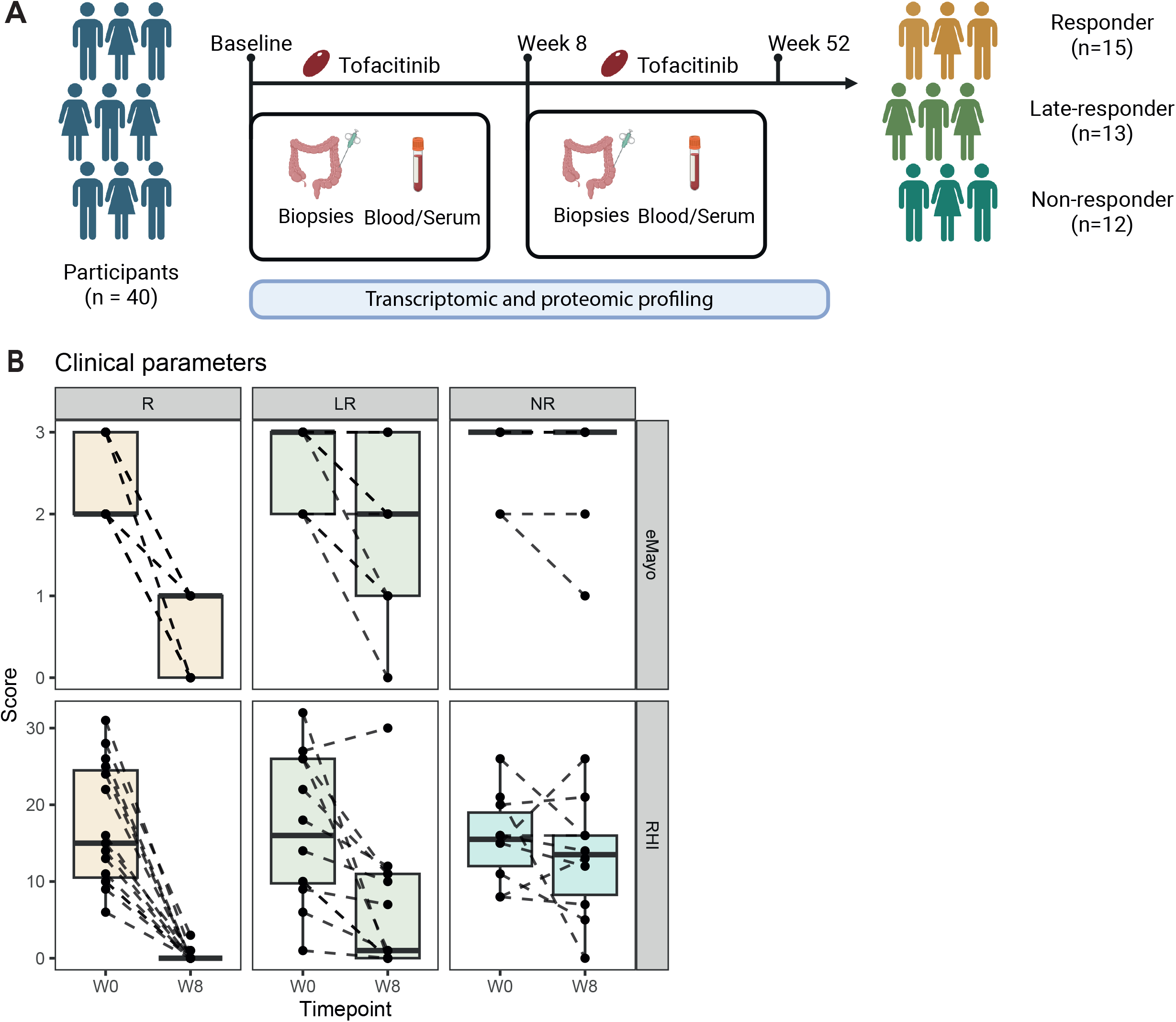
Study design and treatment response to tofacitinib. **(A)** Overview of the cohort and sampling scheme. Forty UC patients scheduled to start tofacitinib were profiled at pretreatment (week 0, W0) and week 8 (W8). Paired inflamed colonic biopsies and blood/serum were collected at both timepoints for transcriptomic and proteomic profiling. Based on clinical assessment through week 52, patients were classified as responders (R, n = 15), late-responders (LR, n = 13) or non-responders (NR, n = 12). **(B)** Clinical parameters by response group and timepoint: endoscopic Mayo subscore (eMayo, top) and Robarts Histopathology Index (RHI, bottom) at W0 and W8, with dashed lines connecting paired samples from the same patient. Colors denote responders (gold), late-responders (green) and non-responders (teal).

### A pro-inflammatory profile marks future responders before treatment initiation

To characterize pretreatment differences, we interrogated the RNA-seq data on biopsies obtained from inflamed colonic lesions prior to the start of treatment. Principal component (PC) analysis revealed no clear separation between future responders and non-responders, suggesting absence of large-scale differential expression **(Figure 2A)**. However, comparative analyses identified 80 response-associated differentially expressed genes (DEGs) pretreatment (**Figure 2B** and **Supplementary Table 1**), where we noted higher baseline expression of genes belonging to the interferon (*STING1*), interleukin 1 (*IL36G, IL36RN*) and), T-cell (*PRKCQ, NFATC2*) related pathways in future responders (**Figure 2C**). Gene set enrichment analyses (GSEA) further corroborated response-associated higher expression of TNF-signaling via NF-κB (p_BH_ = 5.27E-27), immunoregulatory interactions between a lymphoid and a non-lymphoid cell (p_BH_ = 1.93E-22), cytokine-cytokine receptor interaction (p_BH_ = 3.78E-19), FcγR activation (p_BH_ = 2.80E-15), inflammatory response (p_BH_ = 2.57E-14), and signaling by interleukins (p_BH_ = 1.05E-14) (**Figure 2D** and **Supplementary Table 2**).

**Figure 2.**
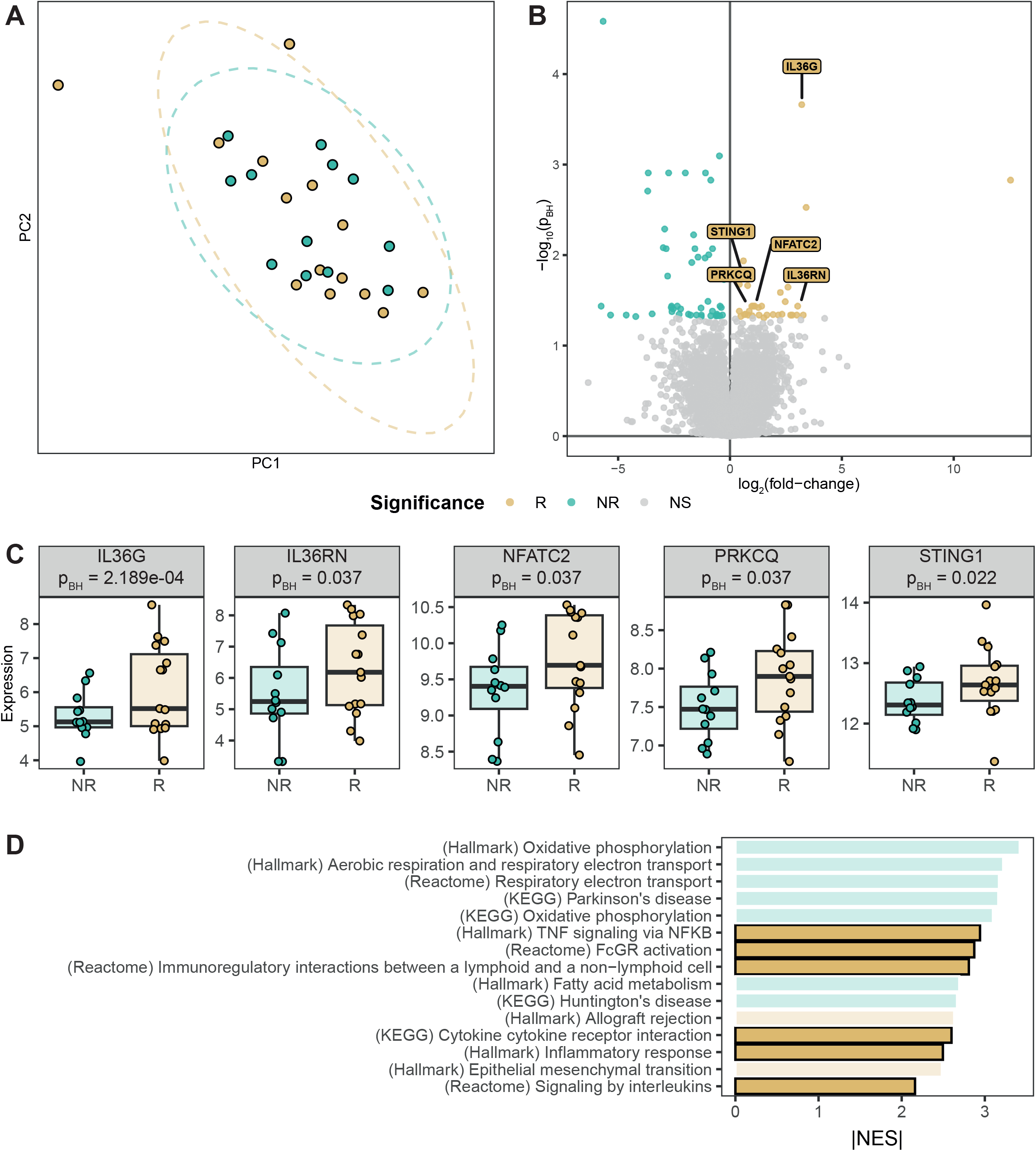
Pretreatment inflammatory signature identifies future responders at week 8. Cross-sectional comparison of inflamed colonic biopsies from future responders (R) versus non-responders (NR) at pretreatment. **(A)** Principal component (PC) analysis of the pretreatment transcriptome, colored by future response: R (gold) and NR (teal). **(B)** Volcano plot of the differential gene expression analysis comparing R with NR at pretreatment. Colored points represent significantly higher expression in R (gold) or NR (teal) at a Benjamini-Hochberg (BH)-adjusted p-value < 0.05; labelled genes are higher in R. **(C)** Variance-stabilized expression of the leading response-associated genes STING1, IL36RN, NFATC2, PRKCQ and IL36G at baseline, grouped by response (NR, R). Reported p-values are BH-adjusted. **(D)** Bar chart of the gene-set enrichment analysis normalized enrichment score (NES), colored by the group in which the set is enriched. Gene sets elevated in future responders include TNF signaling via NF-κB, cytokine-cytokine receptor interaction, FcγR activation, immunoregulatory lymphoid-non-lymphoid interactions and signaling by interleukins. Differential expression: DESeq2, BH-adjusted *p* < 0.05. Enrichment: fgsea, BH-adjusted *p* < 0.05; color, NES. Correlations are Pearson. See Methods for full parameters.

### Responders resolve a JAK-STAT-driven/dependent inflammatory pathway that non-responders retain

Next, we extended our tissue transcriptomic analyses to longitudinal analyses covering samples obtained pretreatment and samples obtained at week 8 to investigate how tofacitinib affected the transcriptome over the course of treatment. PC analysis revealed that responders presented a notable shift over time, which was not observable among non-responders (**Figure 3A**). Longitudinal analyses confirmed the response-associated magnitude of effect, revealing 8,527 DEGs for responders and no DEGs for the non-responders (**Figure 3B** and **Supplementary Tables 3-4**). Comparing the pretreatment response-associated genes longitudinally, we noted that the genes that were significantly higher in responders at baseline were downregulated over time, evidenced by an inverse correlation (r_Pearson_ = −0.46; **Figure 3C**). As expected, responders presented downregulation of pro-inflammatory genes and pathways including JAK-STAT signaling machinery (*JAK1, JAK2, JAK3*), Toll-like receptor signaling (*TLR1, TLR4, TLR5, TLR6, TLR7, TLR8*), MHC class I (*HLA-A, HLA-B, HLA-C, HLA-E, HLA-F*) and II (*HLA-DMA, HLA-DMB, HLA-DRB*), MAPK-signaling (*IL1A, MAP3K8, DUSP4, RELA, FOS, MYC*), as well as the canonical (*IL1B, IL6, CXCL1, CXCL8, ICAM1, MMP9, TNF*) and non-canonical NF-κB signaling (*NFKB2, CD40, CD40LG, TNFRSF8, TNFRSF12A, CD70, LTA*; **Figure 3D-E**). Notably, gene set enrichment analysis indicated that the canonical NF-κB-signaling and MAPK-signaling were downregulated amongst responders but significantly upregulated in non-responders (**Supplementary Table 5-6**). On the other end of the spectrum, drug absorption, distribution, metabolism, and excretion (ADME) were significantly upregulated in responders. Despite the lack of DEGs among non-responders, we noted a correlation among the effect sizes (r_Pearson_ = 0.42; **Figure 3B**), which we could, independent of statistical significance, relate to response-agnostic downregulation of T-cell receptor signaling (*TRAC, CD3D, CD3E, EGR2, ZAP70*) as well as JAK-STAT signaling downstream interferon alpha and gamma response (*IFI44, IFI44L, IFITM1, IFITM2, IFITM3*; **Figure 3E**), indicating that tofacitinib reaches its target regardless of response.

**Figure 3.**
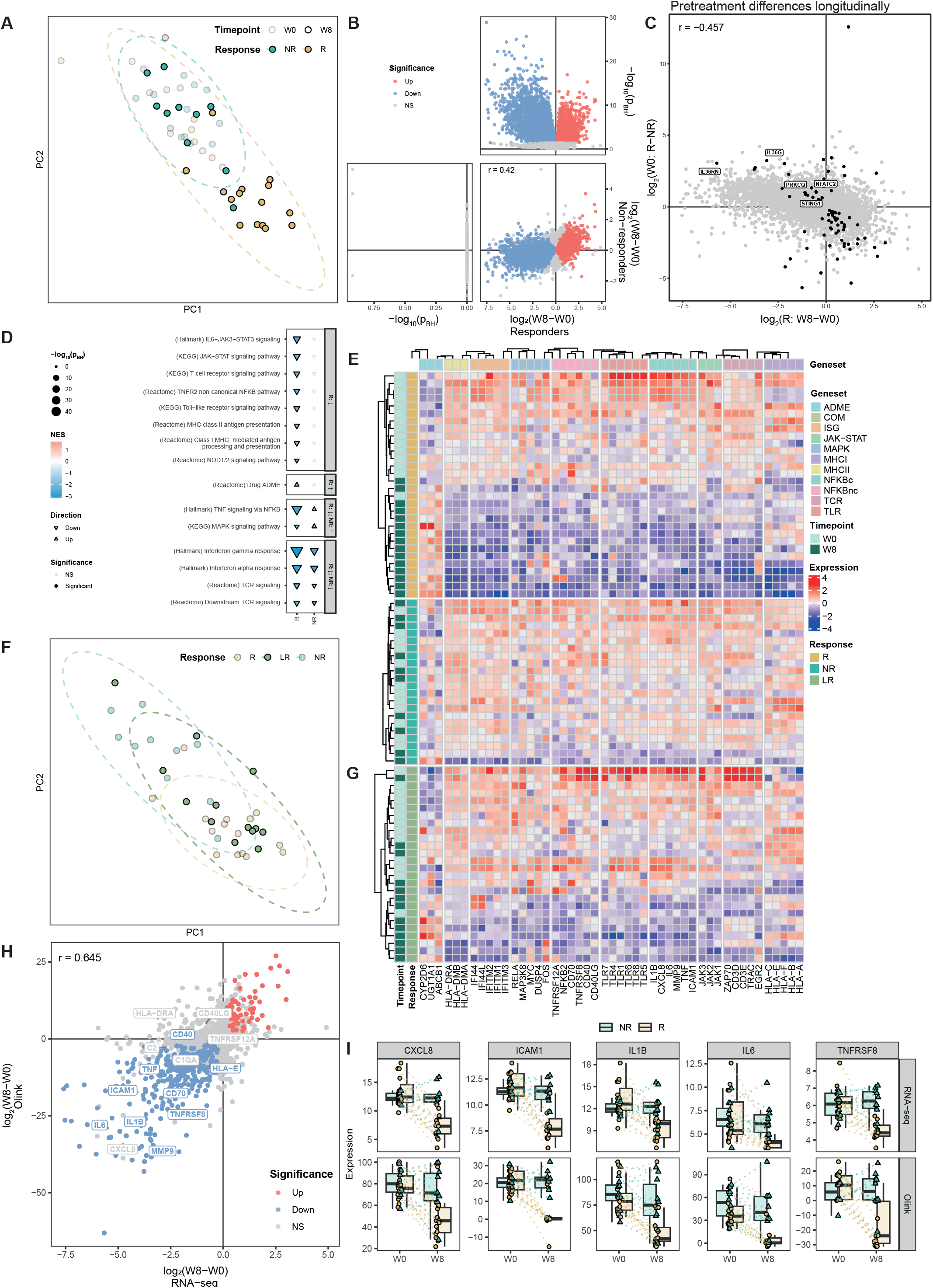
Responder-specific longitudinal resolution of pro-inflammatory gene programs. Longitudinal (week 8 versus week 0) analysis of inflamed biopsies. **(A)** PC analysis colored by response; pretreatment (W0) samples are shown transparent and W8 samples solid. **(B)** Volcano plots of the longitudinal analysis for responders (top right) and non-responders (bottom left), showing effect size as log_2_(fold-change) for week 8 versus week 0 (x-axis) against significance as™log_10_(BH-adjusted p-value) (y-axis). The scatterplot (bottom right) compares effect sizes in R (x-axis) and NR (y-axis), annotated with the Pearson correlation coefficient. **(C)** Scatterplot of longitudinal effect sizes in R (x-axis, log_2(_R: W8™W0)) against the R-versus-NR pretreatment difference (y-axis, log_2(_W0: R™NR)), annotated with the Pearson correlation coefficient. Labelled genes are those highlighted in Figure 2. **(D)** Longitudinal gene-set enrichment analysis of selected pathways (y-axis) for R and NR (x-axis). Triangle orientation denotes direction (up/down), color denotes the NES, and size denotes the ™log_10_(BH-adjusted p-value), and open symbols are non-significant. **(E, G)** Heatmaps of genes illustrative of drug absorption, distribution, metabolism, and excretion (ADME), complement (COM), interferon-stimulated genes (ISG), JAK-STAT, MAPK, MHC class I (MHCI) and class II (MHCII), canonical (NFKBc) and non-canonical (NFKBnc) NF-κB signaling, T-cell receptor (TCR) and Toll-like receptor (TLR) programs across samples, split by **(E)** R and NR and **(G)** LR. Dendrograms were computed using Euclidean distance. **(F)** PC analysis including late-responders at week 8 only. **(H)** Cross-modality comparison of the responder longitudinal differences as log_2(_fold-change) for the transcriptome (x-axis) and proteome (y-axis), annotated with the Pearson correlation coefficient. Colors denote correlated gene-protein pairs. **(I)** Boxplots of representative effectors (CXCL8, ICAM1, IL1B, IL6, TNFRSF8) at week 0 and week 8 in NR and R, stratified by assay (RNA-seq, Olink). Differential expression: DESeq2, BH-adjusted *p* < 0.05. Enrichment: fgsea, BH-adjusted *p* < 0.05; color, NES. Correlations are Pearson. See Methods for full parameters.

We next analyzed the cohort of non-responders that presented HEMI at week 52 to understand whether their transcriptomic profile at week 8 already presented molecular changes similar to the patients that responded at week 8. PC analyses indicated that late responders presented a clustered pattern in between responders and non-responders at week 8 (**Figure 3F**). Indeed, further interrogation of the transcriptome revealed a moderate reduction in the aforementioned inflammatory genes over the initial eight weeks of treatment **(Figure 3G** and **Supplementary Tables 7-8)**.

To understand whether the transcriptomic differences translated into proteomic differences, we utilized a combination of the Cardiometabolic, Inflammation, Neurology and Oncology Olink Explore 384 panels for high-throughput proteomic analyses. Comparing the two modalities revealed a strong correlation between both modalities for responders (**Figure 3H** and **Supplementary Table 9**), but not for non-responders (**Supplementary Figure 1**). This was further exemplified by a cross-modality decrease in both gene and protein expression of NF-κB-signaling genes and proteins IL6, ICAM1, CXCL8, IL1B/IL-1β, and TNFRSF8/CD30 among responders, but not in non-responders (Figure 3I).

### Non-canonical NF-kB downregulation is specific to tofacitinib

To validate our observations, we used a publicly available single-cell RNA-sequencing (scRNA-seq) dataset derived from inflamed colonic biopsies by Melón-Ardanaz *et al*. 2025[10], obtaining data from UC patients starting tofacitinib treatment at week 0 and week 8 from both responders (N = 6) and non-responders (N = 5). Pseudobulk differential gene expression analysis showed a reasonable correlation with our data when contrasting week 8 relative to week 0 for responders (r_Pearson_ = 0.52; **Figure 4A and Supplementary Table 10**), which we could relate to the longitudinal downregulation of the JAK-STAT, interferon alpha and gamma pathways, and NF-κB-signaling and upregulation of drug ADME processes (**Figure 4B** and **Supplementary Table 11**). Of note, non-responders behaved surprisingly different between both cohorts over time: where our own data suggested a non-significant downregulation of JAK-STAT and downstream interferon alpha and gamma pathways, data by Melón-Ardanaz[10] indicated a significant upregulation thereof (**Supplementary Figure 2** and **Supplementary Table 12-13**).

**Figure 4.**
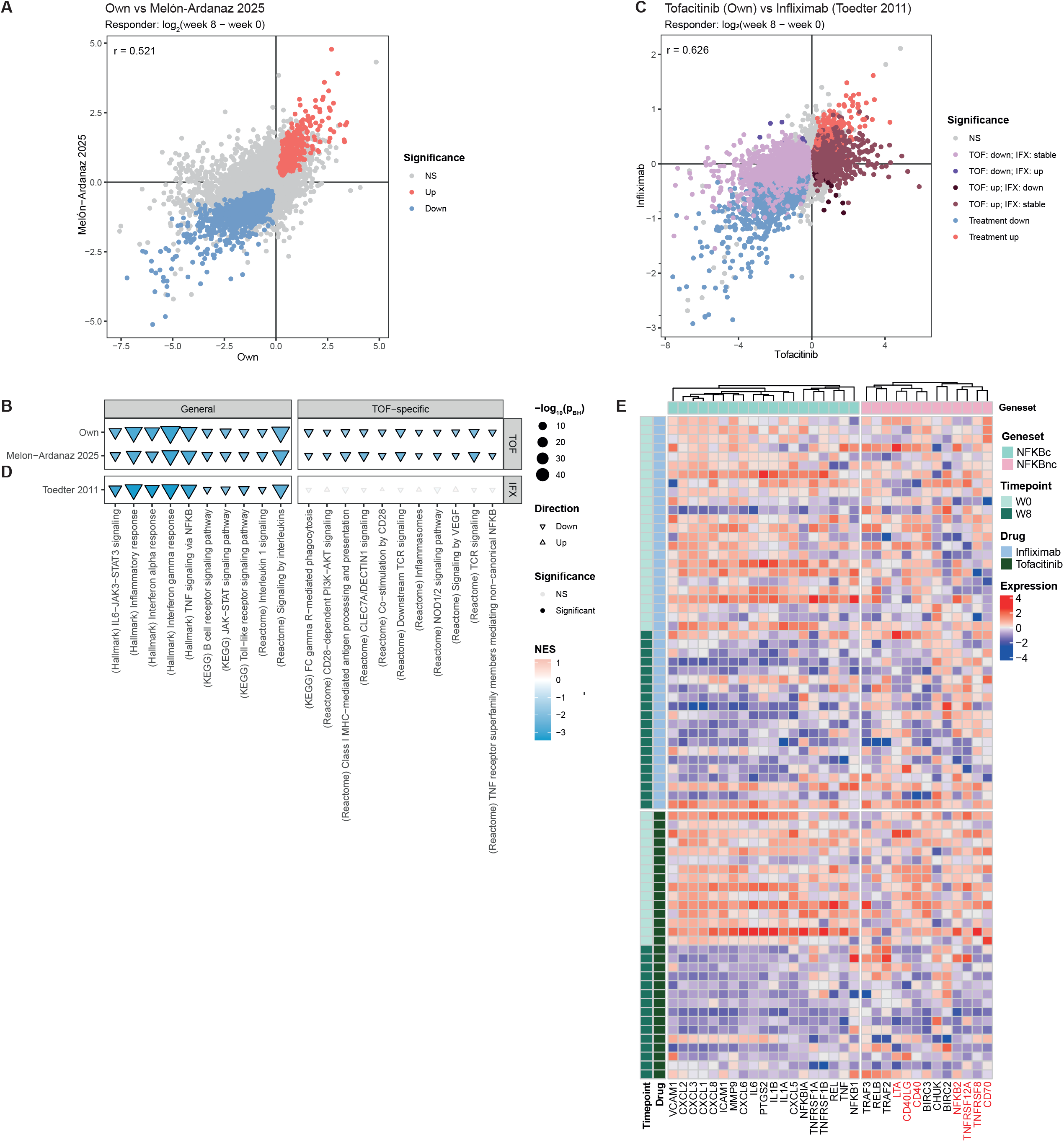
Distinguishing tofacitinib-specific effects from shared inflammatory resolution. **(A)** Scatterplot of the responder longitudinal effect size, log_2(_fold-change) for week 8 versus week 0, from our cohort (Own; x-axis) against the tofacitinib pseudobulk single-cell RNA-sequencing dataset of Melón-Ardanaz *et al*. 2025 (y-axis), annotated with the Pearson correlation coefficient. Colored points are significantly up-(red) or down-regulated (blue) in both cohorts. **(B, D)** Longitudinal gene-set enrichment analysis of selected pathways (x-axis) for tofacitinib responders from **(B)** our cohort and Melón-Ardanaz *et al*. 2025, and **(D)** infliximab responders from Toedter *et al*. 2011. Triangle orientation denotes direction (up/down), color denotes the NES, and size denotes the ™log_10_(BH-adjusted p-value); open symbols are non-significant. Gene sets are faceted into programs shared by tofacitinib and infliximab (General) and those specific to tofacitinib (TOF-specific). (C) Scatterplot of the responder longitudinal effect size, log_2(_fold-change) for week 8 versus week 0, from our cohort (Own; x-axis) against the infliximab microarray dataset of Toedter *et al*. 2011 (y-axis), annotated with the Pearson correlation coefficient. Colors denote concordance class across the two cohorts. TOF, tofacitinib; IFX, infliximab. (E) Heatmap of canonical (NFKBc) and non-canonical (NFKBnc) NF-κB genes across timepoint and drug. Genes highlighted in red are *CD40LG, LTA, TNFRSF8, CD70, TNFRSF12A* and *NFKB2*. Differential expression: DESeq2/limma, BH-adjusted *p* < 0.05. Enrichment: fgsea, BH-adjusted *p* < 0.05; color, NES. Correlations are Pearson. See Methods for full parameters.

To disentangle tofacitinib-specific pathways from general inflammatory resolution, we next interrogated the transcriptomic microarray data from Toedter *et al*. 2011[13], acquiring longitudinal transcriptomic data from inflamed colonic biopsies from UC patients treated for 8 weeks with the TNF inhibitor infliximab, which were categorized as either responders or non-responders. Akin to the earlier observations, differential gene expression analysis revealed a strong correlation between tofacitinib and infliximab over time (r_Pearson_ = 0.63; **Figure 4C** and **Supplementary Table 14**), which was driven by a drug-agnostic downregulation of pro-inflammatory pathways, such as the aforementioned the JAK-STAT signaling and the downstream interferon alpha and gamma response pathways, but also the Toll-like receptor (TLR), interleukin (IL) signaling pathways and their downstream TNF signaling via NF-κB pathways (**Figure 4D** and **Supplementary Table 15**). Focusing on drug-specific behavior, we noted discordant transcriptomic changes for genes associated with the non-canonical NF-κB signaling as well as the related vascular endothelial growth factor (VEGF) signaling, Fc-receptor signaling, inflammasome, TCR pathways, all of which were downregulated in tofacitinib responders but showed little or no change in infliximab responders. We observed a notable separation between the TNFα-driven canonical and TNFα-independent non-canonical NF-κB signaling programs. Canonical NF-κB genes were downregulated over time in both tofacitinib and infliximab responders, consistent with shared resolution of inflammation. In contrast, the non-canonical NF-κB effectors were downregulated in tofacitinib responders while remaining essentially unchanged in infliximab responders (**Figure 4E**). Formal analyses testing the interaction between the response-associated longitudinal differences between both drugs revealed that four of the eight non-canonical NF-κB genes were significantly more downregulated under tofacitinib compared to infliximab: *TNFRSF8* (p_BH_ = 6.58E-11), *CD70* (p_BH_ = 1.34E-05), *CD40* (p_BH_ = 4.60E-05), and *LTA* (p_BH_ = 4.43E-03).

### Shared NF-_k_B pathway pathways reflect distinct gene programs in blood and colonic mucosa

Having established that NF-κB signaling shows tofacitinib-specific behavior over the course of treatment within the colonic mucosa, we sought to understand whether this signature was also observable in peripheral blood (PB). To that end, we extended our RNA-seq to PB. Comparing the responder-associated longitudinal differences over time across mucosal biopsy and PB revealed a weak correlation (r_Pearson_ = 0.05; **Figure 5A** and **Supplementary Table 16**). We nonetheless observed cross-tissue significant downregulation of interferon alpha and gamma signaling, interleukin signaling, and the complement (**Figure 5B** and **Supplementary Table 17**). Notably, both tissue and PB indicated downregulation of the non-canonical NF-κB-signaling pathway. However, we noted that different genes were driving the enrichment signal: in PB, enrichment of the non-canonical NF-κB signaling pathway was primarily driven by the downregulation of proteasome subunits (*PSMA1, PSMB6, PSMD14, PSMC2*), while in the mucosal tissue, enrichment was driven by downregulation of the actual non-canonical NF-κB signaling effectors. Interrogating the core non-canonical NF-κB signaling genes in PB indicated no statistical significance (**Figure 5C**). Notably, PB indicated a significant upregulation of the canonical NF-κB-signaling pathway and Fcγ receptor-mediated phagocytosis, contrasting our earlier observations in mucosal tissue (**Figure 5B**). Further investigation suggested that this pathway-level effect was dominated by immediate-early genes (*IEG; FOS, FOSB, JUN, NR4A1, NR4A2*) and not the core effectors observed in mucosal tissue, which were non-significant in PB (**Figure 5C**). These results indicate that while the canonical and non-canonical NF-κB pathways in mucosal biopsies and PB are enriched, the enrichment occurs through distinct gene programs.

**Figure 5.**
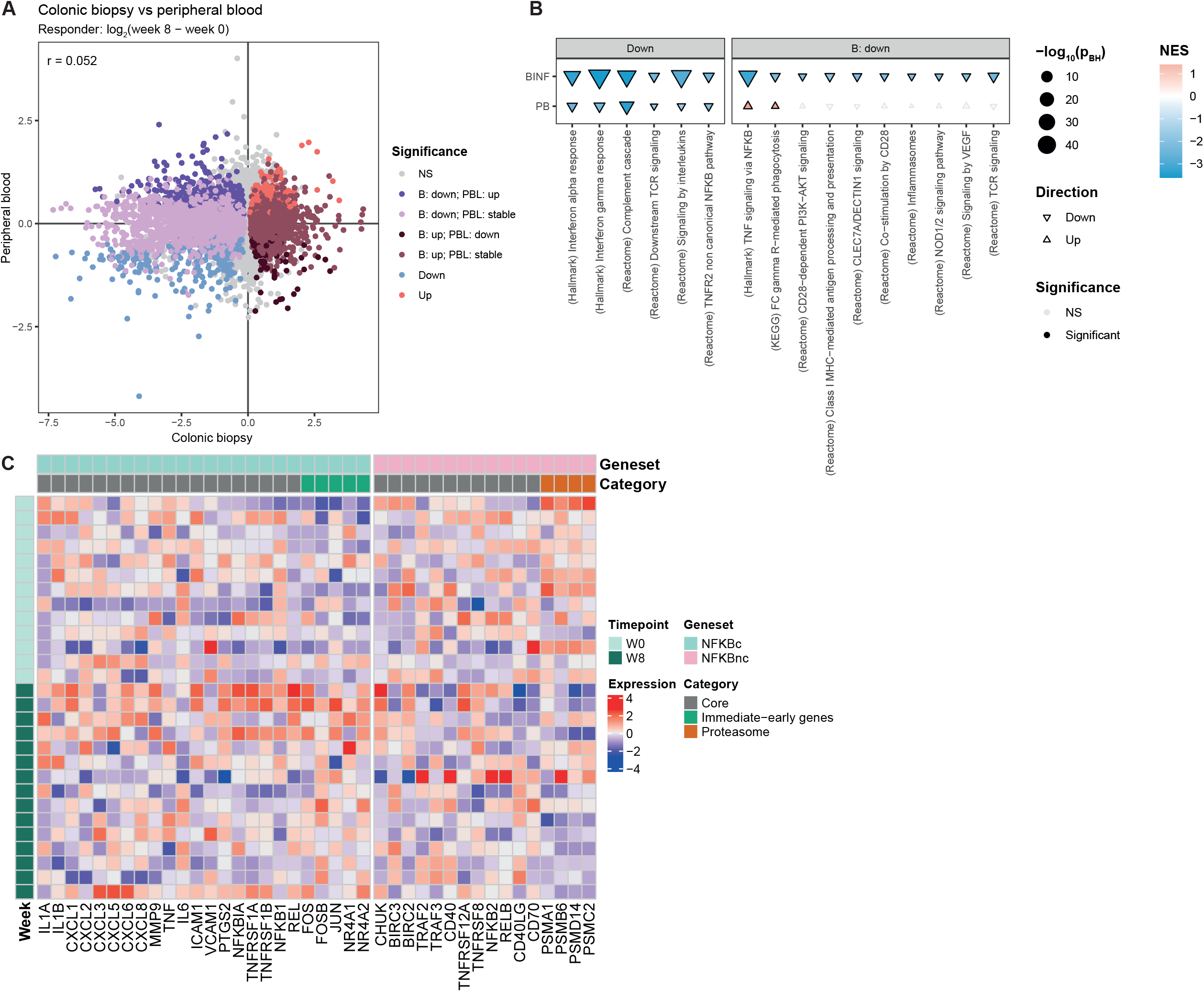
NF-κB pathway concordance between blood and colonic mucosa reflects distinct gene programs. Longitudinal analysis of peripheral blood (PB) in responders. **(A)** Scatterplot of the responder longitudinal effect size, log_2(_fold-change) for week 8 versus week 0, from mucosal biopsies (x-axis) against PB (y-axis), annotated with the Pearson correlation coefficient; colors denote concordance class across the two compartments. **(B)** Longitudinal gene-set enrichment analysis of selected pathways (x-axis) for tofacitinib responders in colonic biopsy (BINF) and PB (y-axis). Triangle orientation denotes direction (up/down), color denotes the NES, and size denotes the ™log_10_(BH-adjusted p-value); open symbols are non-significant. Gene sets are faceted into those downregulated in both compartments (Down) and those downregulated in mucosal biopsy only (B: down). **(C)** Heatmap of canonical (NFKBc) and non-canonical (NFKBnc) NF-κB genes in PB across timepoint, annotated by category: core effectors (Core), immediate-early genes, or proteasome subunits. Differential expression: DESeq2, BH-adjusted *p* < 0.05. Enrichment: fgsea, BH-adjusted *p* < 0.05; colour, NES. Correlations are Pearson. See Methods for full parameters.

### Supervised machine learning analyses identify peripheral blood transcriptomics as most predictive for treatment response

Having profiled multiple molecular modalities across colonic tissue and peripheral blood, we investigated the possibility of predicting future response using samples obtained pretreatment. Given the limited sample size and the lack of an external validation cohort for each modality-tissue combination, we anticipated that our performances may be overfit. We therefore asked which modality-tissue combination best distinguished future responders from non-responders before treatment. Using the TPOT automated machine-learning framework with performance estimated over 100 repeats of stratified five-fold cross-validation, feature selection confined to each training fold. we obtained cross-validated AUROCs of 0.692 (tissue transcriptome), 0.856 (blood transcriptome), 0.550 (serum proteome) and 0.540 (tissue proteome) (**Figure 6**). Only the blood transcriptome separated responders from non-responders clearly above chance, with the tissue transcriptome showing modest separation, and the serum and tissue proteomes performing at or near chance evidenced by the confidence intervals overlapping AUROC = 0.5. Taken together, we identified peripheral blood transcriptomics as the most promising modality for future biomarker studies.

**Figure 6.** Cross-validated pretreatment prediction of tofacitinib response across molecular modalities. Receiver operating characteristic (ROC) curves for supervised classification of responders versus non-responders using pretreatment profiles from four modality-tissue combinations: serum proteome (Olink), tissue proteome (Olink), blood transcriptome (RNA-seq) and tissue transcriptome (RNA-seq).

## DISCUSSION

Here, we present a longitudinal multi-omics characterization of tofacitinib treated patients with active UC, integrating transcriptomic and proteomic profiling across 127 paired mucosal and peripheral-blood samples providing the largest and most comprehensive multi-omic dataset to date on tofacitinib response. We confirm the clinical phenotype of inflammatory resolution in responders at both the transcriptomic and proteomic level, manifesting as a significant reduction in pro-inflammatory pathways, including IL-6/JAK-STAT3 signaling and JAK-STAT signaling more broadly[14, 15, 16, 17]. This general transcriptomic downregulation in responders was validated against earlier data from Melón-Ardanaz *et al*. 2025[10]. While the reduction in inflammatory signaling is observable in attenuated form in late responders, practically no such reduction is observable in non-responders, except for the JAK-STAT-downstream interferon-α and −γ pathways, corroborating tofacitinib’s engagement with its target even in non-responders.

Comparison of responders and non-responders before treatment showed that the discriminating genes represented a specific elevation of the effector programs that propagate through tofacitinib’s molecular target, namely interferon, IL-6-family, IL-2/common-γ-chain and TCR-driven signaling, all significantly higher at baseline in future responders. Notably, this effector program would subsequently collapse during tofacitinib treatment. *STING1* sits upstream of the type I interferon-JAK1 axis[18], and *PRKCQ* and *NFATC2* are part of the TCR signaling that generates the γ-chain cytokines whose transduction is JAK1/JAK3-dependent[19]. Elevated expression of these genes at baseline therefore suggests an interferon- and T-cell-primed mucosa that is sensitive to tofacitinib. By contrast, the elevated IL-36 components (the agonist IL36G and its antagonist IL36RN) signal through the MyD88-NF-κB axis rather than JAK[20]. Despite this JAK-independence, the longitudinal resolution of NF-κB signaling under tofacitinib points to an indirect effect on this pathway. Notably, Melón-Ardanaz *et al*. 2025[10] reported NF-κB signaling to be elevated in non-responders, contrasting our own observations. Our analysis of their data indicates that while their responders behave similarly to ours, their non-responders diverge markedly, aggravating their pro-inflammatory phenotype at the transcriptomic level. Although the basis for this discrepancy is unclear, an actively worsening non-responder phenotype is consistent with non-responder inflammation being driven through pathways that JAK inhibition does not resolve. Response to tofacitinib is dependent on inflammation being by the JAK-STAT pathway, which appears to be elevated at baseline. By contrast, non-responders may represent a distinct endotype in which inflammation is sustained through JAK-independent or stromal pathways that JAK inhibition cannot ameliorate.

Comparing tofacitinib with infliximab clarified which of the aforementioned changes are drug-specific. Downregulation of JAK-STAT and interferon signaling was observable in both tofacitinib- and infliximab-treated patients and occurred largely independently of response, indicating general resolution of inflammation rather than a tofacitinib-specific mechanism. The tofacitinib-distinctive signal instead resided in the non-canonical NF-κB pathway: whereas canonical NF-κB effectors (*IL1B, IL6, CXCL8, ICAM1, TNF*) fell with both drugs, the non-canonical effectors (*NFKB2, CD70, LTA, CD40LG, TNFRSF8, TNFRSF12A*) were downregulated by tofacitinib but unchanged by infliximab. This is mechanistically consistent with infliximab’s inhibition of TNFα thereby abrogating the canonical TNF-driven NF-κB signaling pathway. Tofacitinib, by contrast, may reach the non-canonical arm indirectly through depletion of the JAK1/JAK3-dependent T cell-derived ligands (CD40L, CD70, LTA) that sustain non-canonical NF-κB activation in stromal and myeloid compartments [21, 22].

Extending the analysis to peripheral blood revealed compartment-specific features. Although several pathway labels were shared between mucosa and peripheral blood, the underlying genes driving the enrichment differed. Where tissue presented downregulation of the core canonical and non-canonical NF-κB signaling genes, in peripheral blood, non-canonical NF-κB enrichment was driven by proteasome subunits and canonical NF-κB enrichment reflected immediate-early genes (*FOS, FOSB, JUN, NR4A1, NR4A2*). Pathway-level concordance between blood and mucosa therefore does not imply a shared biological process.

Beyond characterizing the mechanism of response, we investigated whether pretreatment molecular profiles could predict treatment response to tofacitinib. Across four modality-tissue combinations, only the peripheral blood transcriptome separated future responders from non-responders appreciably above chance, outperforming colonic tissue transcriptomics and both proteomic modalities in this cohort. We acknowledge that the magnitude of the blood-transcriptome performance is implausibly high, suggesting overfitting as we did not have access to an external cohort for all modality-tissue combinations. While repeated cross-validation reduces variance, it does not eliminate bias when the feature space vastly exceeds the sample size, and automated pipeline selection compounds it [23, 24]. Confirmation will require an independent, adequately powered cohort with a locked analytical pipeline. We therefore present this as hypothesis-generating: the blood transcriptome is the modality most worth pursuing, corroborating our earlier DNA methylome study [9]. We acknowledge that the predictive signal seemingly contrasts our earlier observations that peripheral blood does not serve as a straightforward proxy for the mucosal response as we indicated that the shared pathways between compartments were driven by different genes. We argue that a peripheral signature need not mirror the mucosal effector program to be discriminative and may instead reflect a systemic immune state that associates with a patient’s likelihood of mucosal response without being its mechanistic driver. Clinically, the utility of peripheral blood over mucosal biopsy is evident, with the former representing a minimally invasive sample that can be more readily acquired compared to the latter.

A particular strength of this study is the blinded central reading of endoscopic and histologic assessments, enabling stringent phenotypic classification that underpins the transcriptomic and proteomic contrasts reported here. Several limitations should also be acknowledged. First, we used data from Melón-Ardanaz *et al*.[10] and Toedter *et al*.[13], who maintained different response criteria and experimental setups (scRNA-seq versus transcriptomics microarray). Second, the response-discriminating biology is expressed predominantly at the pathway level rather than at the single-gene level, as few baseline genes survived individual correction; this suggests that a clinical assay will require multiple genes for accurate prediction. A corollary is that tofacitinib response cannot be reduced to a single dimension and represents a complex phenotype. Third, the baseline analyses are cross-sectional and associative, and cannot establish that the pretreatment state causes response. Fourth, bulk transcriptomics cannot separate reduced per-cell signaling from shifts in the abundance of the cells that carry it, which would require complementary single-cell analyses, as presented in Melón-Ardanaz *et al*. 2025[10]. However, single-cell analyses remain expensive and experimentally demanding, limiting their scalability for clinical use.

In conclusion, our data support a model where tofacitinib response in UC depends on a JAK-dependent pro-inflammatory phenotype that tofacitinib can dismantle, in contrast to non-responders representing distinct, JAK-independent pathways. Our findings provide a foundation for treatment response studies in UC towards the development and validation of predictive biomarkers in colonic mucosa and blood, which could ultimately result in more personalized treatment strategies.

## Competing interests

IF: None

DJ: None

KL: None

ALY: Consultancy/lecture fees from Janssen, Johnson & Johnson, DeciBio. Stock options from GlaxoSmithKline and Haleon. Grants received from Crohn’s and Colitis Foundation.

SG: None

RR: None

EC: None

JJ: None

KG: Research grants from AbbVie, Eli Lilly, Pfizer Inc, Celltrion and Galapagos; consultancy fees from AbbVie, Galapagos, Gilead, Janssen Pharmaceuticals, Pfizer Inc., and Takeda and speaker’s honoraria from AbbVie, Eli Lilly, Ferring, Galapagos, Janssen Pharmaceuticals, Pfizer Inc, Takeda and Tillotts.

ML: Consultancy/lecture fees from Abbvie, Bristol Myers Squibb, Eli Lilly, Alfasigma, Janssen-Cilag, Johnson & Johnson, Medtronic, Pfizer, Takeda, Tillotts. These fees are invoiced by the speaker fee desk of the university. Grants received from ZonMW, Alfasigma, NFU transformation deal and TKI. AM: None

WJ: Research grants from RB, Friesland Campina, Bristol Myers Squibb, Alimentiv, Acelabio, MRM Health, Trained Therapeutics Discovery BV. Consultant to Liminal Labs BV.

GD: Research grants from Abbvie, Alimentiv, Bristol Myers Squibb, Johnson & Johnson, Pfizer, Takeda. Consulting fees from Abbvie, Agomab, Alimentiv, AstraZeneca, AMT, Bristol Myers Squibb, Boehringer Ingelheim, Celltrion, Eli Lilly, Exeliom Biosciences, Galapagos, Index Pharmaceuticals, Kaleido, GlaxoSmithKline, Gossamerbio, Pfizer, Immunic, Johnson and Johnson, Polpharma, Procise Diagnostics, Prometheus laboratories, Prometheus Biosciences, Progenity, Protagonist, Ventyx. Speaker’s fees from Abbvie, Arena, Boehringer Ingelheim, Celltrion, Galapagos, Gilead, Pfizer, BMS, Takeda. Data monitoring board: Galapagos, AstraZeneca, Seres Health.

## Funding

This study was sponsored by the Amsterdam UMC and Pfizer. Pfizer was not involved in the study design, data collection, analysis, and interpretation of data.

## Author contributions

DJ, IF, SG, GD, AM, and WJ were involved in the study concept and design. DJ, IF, SG, RR, EC, JJ, KG, ML, AM, and GD were involved in data acquisition. SG performed all statistical analyses. DJ, IF, KL, ALY, SG, RR, AM, and GD were involved in data analysis and interpretation of data. DJ, IF, KL, ALY and GD drafted the initial manuscript. Critical revision of the final manuscript for important intellectual content was performed by all authors.

## Supplementary figures

**Supplementary Figure 1**. Cross-sectional proteomic comparison of inflamed colonic biopsies from future responders (R) versus non-responders (NR) at pretreatment. Volcano plots of the longitudinal analysis for responders (top right) and non-responders (bottom left), showing effect size as log_2(_fold-change) for week 8 versus week 0 (x-axis) against significance as ™log_10_(BH-adjusted p-value) (y-axis). The scatterplot (bottom right) compares effect sizes in R (x-axis) and NR (y-axis), annotated with the Pearson correlation coefficient.

**Supplementary Figure 2**. Comparison of inflamed colonic biopsies from future responders (R) versus non-responders (NR) at pretreatment. **(A, B)** Scatterplot of the differential **(A)** gene expression and (B) gene set enrichment analysis effect size, log_2(_fold-change) and NES gene and gene set enrichment analyses, respectively, from our cohort (Own; x-axis) against the tofacitinib pseudobulk single-cell RNA-sequencing dataset of Melón-Ardanaz *et al*. 2025 (y-axis), annotated with the Pearson correlation coefficient. Colored points indicate significance and direction in both cohorts. **(C)** Longitudinal gene-set enrichment analysis of selected pathways (x-axis) for tofacitinib responders and non-responders from our cohort and Melón-Ardanaz *et al*. 2025.

## Supplementary tables

**Supplementary Table 1**. Differential gene expression analysis results from inflamed mucosal biopsy comparing responders with non-responders at baseline, derived using DESeq2. Columns: gene symbol; Ensembl identifier; baseMean, the mean normalized count across all samples; log2FoldChange; lfcSE, standard error of the log2 fold change; stat, Wald statistic; pvalue, nominal p-value; padj, Benjamini-Hochberg adjusted p-value.

**Supplementary Table 2**. Gene set enrichment analysis results from inflamed mucosal biopsy comparing responders with non-responders at baseline, derived using pre-ranked fgsea against the Wald statistic obtained from Supplementary Table 1. Columns: pathway, pathway identifier; pvalue, nominal p-value; padj, Benjamini-Hochberg adjusted p-value; log2err, the expected error for the standard deviation of the p-value logarithm; ES, enrichment score; NES, enrichment score normalized to mean enrichment of random samples of the same size; size, size of the pathway; leadingEdge; genes that drive the enrichment.

**Supplementary Table 3**. Differential gene expression analysis results from inflamed mucosal biopsy comparing week 8 against baseline for responders, derived using DESeq2. Columns: gene symbol; Ensembl identifier; baseMean, the mean normalized count across all samples; log2FoldChange; lfcSE, standard error of the log2 fold change; stat, Wald statistic; pvalue, nominal p-value; padj, Benjamini-Hochberg adjusted p-value.

**Supplementary Table 4**. Gene set enrichment analysis results from inflamed mucosal biopsy comparing week 8 against baseline for responders, derived using pre-ranked fgsea against the Wald statistic obtained from Supplementary Table 3. Columns: pathway, pathway identifier; pvalue, nominal p-value; padj, Benjamini-Hochberg adjusted p-value; log2err, the expected error for the standard deviation of the p-value logarithm; ES, enrichment score; NES, enrichment score normalized to mean enrichment of random samples of the same size; size, size of the pathway; leadingEdge; genes that drive the enrichment.

**Supplementary Table 5**. Differential gene expression analysis results from inflamed mucosal biopsy comparing week 8 against baseline for non-responders, derived using DESeq2. Columns: gene symbol; Ensembl identifier; baseMean, the mean normalized count across all samples; log2FoldChange; lfcSE, standard error of the log2 fold change; stat, Wald statistic; pvalue, nominal p-value; padj, Benjamini-Hochberg adjusted p-value.

**Supplementary Table 6**. Gene set enrichment analysis results from inflamed mucosal biopsy comparing week 8 against baseline for non-responders, derived using pre-ranked fgsea against the Wald statistic obtained from Supplementary Table 5. Columns: pathway, pathway identifier; pvalue, nominal p-value; padj, Benjamini-Hochberg adjusted p-value; log2err, the expected error for the standard deviation of the p-value logarithm; ES, enrichment score; NES, enrichment score normalized to mean enrichment of random samples of the same size; size, size of the pathway; leadingEdge; genes that drive the enrichment.

**Supplementary Table 7**. Differential gene expression analysis results from inflamed mucosal biopsy comparing week 8 against baseline for late responders, derived using DESeq2. Columns: gene symbol; Ensembl identifier; baseMean, the mean normalized count across all samples; log2FoldChange; lfcSE, standard error of the log2 fold change; stat, Wald statistic; pvalue, nominal p-value; padj, Benjamini-Hochberg adjusted p-value.

**Supplementary Table 8**. Gene set enrichment analysis results from inflamed mucosal biopsy comparing week 8 against baseline for late responders, derived using pre-ranked fgsea against the Wald statistic obtained from Supplementary Table 7. Columns: pathway, pathway identifier; pvalue, nominal p-value; padj, Benjamini-Hochberg adjusted p-value; log2err, the expected error for the standard deviation of the p-value logarithm; ES, enrichment score; NES, enrichment score normalized to mean enrichment of random samples of the same size; size, size of the pathway; leadingEdge; genes that drive the enrichment.

**Supplementary Table 9**. Differential protein expression analysis results from inflamed mucosal biopsy on combined Cardiometabolic, Inflammation, Neurology and Oncology Olink Explore 384 data comparing week 8 against baseline for responders, derived using limma. Columns: protein symbol; AveExpr, the average expression across all samples; log2FoldChange; stat, t-statistic; pvalue, nominal p-value; padj, Benjamini-Hochberg adjusted p-value; OlinkID; Olink identifier; UniProt, UniProt identifier; Panel, source of the probe; Panel_Lot_Nr, panel lot number.

**Supplementary Table 10**. Differential gene expression analysis results from mucosal biopsy on pseudobulked single-cell RNA-sequencing data obtained from GSE253006 (Mélon-Ardanaz 2025) comparing week 8 against baseline for responders, derived using DESeq2. Columns: gene symbol; Ensembl identifier; baseMean, the mean normalized count across all samples; log2FoldChange; lfcSE, standard error of the log2 fold change; stat, Wald statistic; pvalue, nominal p-value; padj, Benjamini-Hochberg adjusted p-value.

**Supplementary Table 11**. Gene set enrichment analysis results from mucosal biopsy on pseudobulked single-cell RNA-sequencing data obtained from GSE253006 (Mélon-Ardanaz 2025) comparing week 8 against baseline for responders, derived using pre-ranked fgsea against the Wald statistic obtained from Supplementary Table 9. Columns: pathway, pathway identifier; pvalue, nominal p-value; padj, Benjamini-Hochberg adjusted p-value; log2err, the expected error for the standard deviation of the p-value logarithm; ES, enrichment score; NES, enrichment score normalized to mean enrichment of random samples of the same size; size, size of the pathway; leadingEdge; genes that drive the enrichment.

**Supplementary Table 12**. Differential gene expression analysis results from mucosal biopsy on pseudobulked tofacitinib single-cell RNA-sequencing data obtained from GSE253006 (Mélon-Ardanaz 2025) comparing week 8 against baseline for non-responders, derived using DESeq2. Columns: gene symbol; Ensembl identifier; baseMean, the mean normalized count across all samples; log2FoldChange; lfcSE, standard error of the log2 fold change; stat, Wald statistic; pvalue, nominal p-value; padj, Benjamini-Hochberg adjusted p-value.

**Supplementary Table 13**. Gene set enrichment analysis results from mucosal biopsy on pseudobulked tofacitinib single-cell RNA-sequencing data obtained from GSE253006 (Mélon-Ardanaz 2025) comparing week 8 against baseline for non-responders, derived using pre-ranked fgsea against the Wald statistic obtained from Supplementary Table 11. Columns: pathway, pathway identifier; pvalue, nominal p-value; padj, Benjamini-Hochberg adjusted p-value; log2err, the expected error for the standard deviation of the p-value logarithm; ES, enrichment score; NES, enrichment score normalized to mean enrichment of random samples of the same size; size, size of the pathway; leadingEdge; genes that drive the enrichment.

**Supplementary Table 14**. Differential gene expression analysis results from mucosal biopsy on infliximab microarray data obtained from GSE23597 (Toedter 2011) comparing week 8 against baseline for responders, derived using limma. Columns: gene symbol; AveExpr, the average expression across all samples; log2FoldChange; stat, t-statistic; pvalue, nominal p-value; padj, Benjamini-Hochberg adjusted p-value; ProbeID; Affymetrix Human Genome U133 Plus 2.0 probe identifier.

**Supplementary Table 15**. Gene set enrichment analysis results from mucosal biopsy on infliximab microarray data obtained from GSE23597 (Toedter 2011) comparing week 8 against baseline for responders, derived using pre-ranked fgsea against the Wald statistic obtained from Supplementary Table 13. Columns: pathway, pathway identifier; pvalue, nominal p-value; padj, Benjamini-Hochberg adjusted p-value; log2err, the expected error for the standard deviation of the p-value logarithm; ES, enrichment score; NES, enrichment score normalized to mean enrichment of random samples of the same size; size, size of the pathway; leadingEdge, genes that drive the enrichment.

**Supplementary Table 16**. Differential gene expression analysis results from peripheral blood comparing week 8 against baseline for responders, derived using DESeq2. Columns: gene symbol; Ensembl identifier; baseMean, the mean normalized count across all samples; log2FoldChange; lfcSE, standard error of the log2 fold change; stat, Wald statistic; pvalue, nominal p-value; padj, Benjamini-Hochberg adjusted p-value.

**Supplementary Table 17**. Gene set enrichment analysis results from peripheral blood comparing week 8 against baseline for responders, derived using pre-ranked fgsea against the Wald statistic obtained from Supplementary Table 15. Columns: pathway, pathway identifier; pvalue, nominal p-value; padj, Benjamini-Hochberg adjusted p-value; log2err, the expected error for the standard deviation of the p-value logarithm; ES, enrichment score; NES, enrichment score normalized to mean enrichment of random samples of the same size; size, size of the pathway; leadingEdge; genes that drive the enrichment.

## Notes

### Clinical Trial

NL-OMON47601

### Author Declarations

Medical Ethics Committee of the Academic Medical Center gave ethical approval for this work under reference METC2017_120.

## REFERENCES

1 Scott LJ. Tofacitinib: a review of its use in adult patients with rheumatoid arthritis. Drugs 2013;73:857–74.

2 Sandborn WJ, Ghosh S, Panes J, Vranic I, Su C, Rousell S, et al. Tofacitinib, an oral Janus kinase inhibitor, in active ulcerative colitis. N Engl J Med 2012;367:616–24.

3 De Vries LCS, Wildenberg ME, De Jonge WJ, D’Haens GR. The Future of Janus Kinase Inhibitors in Inflammatory Bowel Disease. J Crohns Colitis 2017;11:885–93.

4 Sandborn WJ, Su C, Sands BE, D’Haens GR, Vermeire S, Schreiber S, et al. Tofacitinib as Induction and Maintenance Therapy for Ulcerative Colitis. N Engl J Med 2017;376:1723–36.

5 Taxonera C, Olivares D, Alba C. Real-World Effectiveness and Safety of Tofacitinib in Patients With Ulcerative Colitis: Systematic Review With Meta-Analysis. Inflamm Bowel Dis 2022;28:32–40.

6 Steenholdt C, Dige Ovesen P, Brynskov J, Seidelin JB. Tofacitinib for Acute Severe Ulcerative Colitis: A Systematic Review. J Crohns Colitis 2023;17:1354–63.

7 Long MD, Afzali A, Fischer M, Hudesman D, Abdalla M, McCabe R, et al. Tofacitinib Response in Ulcerative Colitis (TOUR): Early Response After Initiation of Tofacitinib Therapy in a Real-world Setting. Inflamm Bowel Dis 2023;29:570–8.

8 Lin CH, Liu WS, Wan C, Wang HH. Effectiveness of tofacitinib in patients with ulcerative colitis: an updated systematic review and meta-analysis of real-world studies. BMJ Open Gastroenterol 2024;11.

9 Joustra V, Li Yim AYF, van Gennep S, Hageman I, de Waard T, Levin E, et al. Peripheral Blood DNA Methylation Signatures and Response to Tofacitinib in Moderate-to-severe Ulcerative Colitis. J Crohns Colitis 2024;18:1179–89.

10 Melón-Ardanaz E, Veny M, Corraliza AM, Gudiño V, Garrido-Trigo A, Sanzo-Machuca Á, et al. Differential effects of tofacitinib on macrophage activation contribute to lack of response in ulcerative colitis patients. J Crohns Colitis 2025;19.

11 Mukherjee A, Tsuchiwata S, Nicholas T, Cook JA, Modesto I, Su C, et al. Exposure-Response Characterization of Tofacitinib Efficacy in Moderate to Severe Ulcerative Colitis: Results From Phase II and Phase III Induction and Maintenance Studies. Clin Pharmacol Ther 2022;112:90–100.

12 van Gennep S, Fung ICN, Jong DC, Ramkisoen RK, Clasquin E, de Jong J, et al. Histological Outcomes and JAK-STAT Signalling in Ulcerative Colitis Patients Treated with Tofacitinib. J Crohns Colitis 2024;18:1283–91.

13 Toedter G, Li K, Marano C, Ma K, Sague S, Huang CC, et al. Gene expression profiling and response signatures associated with differential responses to infliximab treatment in ulcerative colitis. Am J Gastroenterol 2011;106:1272–80.

14 Veltkamp SHC, Voorneveld PW. The Cell-Specific Effects of JAK1 Inhibitors in Ulcerative Colitis. J Clin Med 2025;14.

15 Barnhoorn MC, van der Meulen-de Jong AE, Schrama E, Plug LG, Verspaget HW, Fibbe WE, et al. Cytokine Mixtures Mimicking the Local Milieu in Patients with Inflammatory Bowel Disease Impact Phenotype and Function of Mesenchymal Stromal Cells. Stem Cells Transl Med 2022;11:932–45.

16 Iboshi Y, Nakamura K, Fukaura K, Iwasa T, Ogino H, Sumida Y, et al. Increased IL-17A/IL-17F expression ratio represents the key mucosal T helper/regulatory cell-related gene signature paralleling disease activity in ulcerative colitis. J Gastroenterol 2017;52:315–26.

17 Salas A, Hernandez-Rocha C, Duijvestein M, Faubion W, McGovern D, Vermeire S, et al. JAK-STAT pathway targeting for the treatment of inflammatory bowel disease. Nat Rev Gastroenterol Hepatol 2020;17:323–37.

18 Decout A, Katz JD, Venkatraman S, Ablasser A. The cGAS-STING pathway as a therapeutic target in inflammatory diseases. Nat Rev Immunol 2021;21:548–69.

19 Morley SC, Weber KS, Kao H, Allen PM. Protein kinase C-theta is required for efficient positive selection. J Immunol 2008;181:4696–708.

20 Yuan ZC, Xu WD, Liu XY, Liu XY, Huang AF, Su LC. Biology of IL-36 Signaling and Its Role in Systemic Inflammatory Diseases. Front Immunol 2019;10:2532.

21 Rao DA, Gurish MF, Marshall JL, Slowikowski K, Fonseka CY, Liu Y, et al. Pathologically expanded peripheral T helper cell subset drives B cells in rheumatoid arthritis. Nature 2017;542:110–4.

22 Crotty S. T Follicular Helper Cell Biology: A Decade of Discovery and Diseases. Immunity 2019;50:1132–48.

23 Vabalas A, Gowen E, Poliakoff E, Casson AJ. Machine learning algorithm validation with a limited sample size. PLoS One 2019;14:e0224365.

24 Cawley GC, Talbot NLC. On Over-fitting in Model Selection and Subsequent Selection Bias in Performance Evaluation. Journal of Machine Learning Research 2010;11:2079–107.

