## Supplementary Information for "Longitudinal multi-omics profiling of tofacitinib response identifies a pre-existing Janus kinase-tractable inflammatory state in ulcerative colitis"

#### Transcriptomic analysis

RNA from peripheral blood was isolated using the PAXgene blood RNA kit (Qiagen), whereafter globin was depleted using the GLOBINclear kit (Thermo Fisher). RNA from colonic biopsies were isolated using the RNeasy extraction kit (Qiagen). Paired-end bulk RNA-sequencing (RNA-seq; 100bp) was conducted by Covance, Labcorp to a depth of 50M reads per sample. Raw sequencing reads were assessed for quality using FastQC (v0.11.9) [1]. Reads were aligned to the human reference genome (GRCh38) using STAR aligner (v2.7.10a) [2]. Gene-level read counts were obtained using featureCounts from the Subread package (v2.0.1) using Gencode (v38) for gene models[3]. Differential gene expression analysis was performed using DESeq2 (v1.42.0)[4]. For pretreatment responder versus non-responders analyses, models were adjusted for age, sex, baseline concomitant prednisone use, anti-TNF exposure, vedolizumab exposure, and eMayo subscore. For longitudinal analyses, paired samples were included in the design formula.

Differentially expressed genes (DEGs) were defined as those with a Benjamini-Hochberg-adjusted p < 0.05. Gene set enrichment analysis (GSEA) was performed using fgsea (v1.28.0)[5] on genes ranked by the Wald statistic (DESeq2) or the moderated t-statistic (limma), against the Hallmark, KEGG, and Reactome collections obtained from the Molecular Signatures Database (MSigDB v2023.2.Hs). Pathways with a Benjamini-Hochberg-adjusted p < 0.05 were considered significantly enriched. The normalized enrichment score (NES) denotes the direction and magnitude of enrichment.

Drug-by-time interactions were estimated for each gene as the difference between the tofacitinib and infliximab responder log₂ fold-changes (week 8 versus week 0), with variance equal to the sum of the per-drug squared standard errors. Interaction effects were tested by a two-sided Wald test and corrected for multiple testing using the Benjamini-Hochberg procedure. Where multiple microarray probes mapped to a gene, the probe with the highest average expression was retained. This contrast was applied to the *a priori* non-canonical NF-κB gene set.

#### Proteomic analysis

Proteomic profiling was performed using the Olink Explore 384 assay combining the Cardiometabolic, Inflammation, Neurology and Oncology panels (Olink Proteomics AB, Uppsala, Sweden) yielding protein expression profiles of roughly 1500 proteins. Probes measuring the same proteins were summarized by the mean. Differential protein expression analysis of Olink NPX values was performed using limma (v3.58.1)[6]. Baseline predictive analyses were adjusted for baseline anti-TNF exposure, baseline eMayo score, and concomitant prednisone use. Longitudinal analyses additionally accounted for paired samples. Differentially expressed proteins (DEPs) were defined as those with a Benjamini-Hochberg-adjusted p < 0.05.

#### Public data analysis

Publicly available processed tofacitinib single-cell RNA-sequencing (scRNA-seq) from GSE253006[7] and infliximab bulk transcriptomic microarray data from GSE23597[8] were acquired for validation and specificity purposes, respectively. The tofacitinib scRNA-seq data pertained a Chromium 10X 3’ gene expression experiment performed on mucosal biopsy samples obtained from responders (n = 6) and non-responders (n = 5) at week 0 and week 8, treated with 10 mg tofacitinib BID. Response was defined as a decrease in eMayo subscore ≥ 1. Data were imported into R using Seurat, pseudobulked by summing the reads per genes per sample and then subjected to differential gene expression analysis using DESeq2. The infliximab transcriptomic microarray pertained an Affymetrix Human Genome U133 Plus 2.0 Array gene expression experiment performed on mucosal biopsy samples obtained from responders (n = 25) and non-responders (n = 7) obtained at week 0 and week 8, treated with either 5 mg or 10 mg infliximab. Response was defined as a decrease in total Mayo score ≥ 3, representing at least 30% in combination with a decrease in rectal bleeding subscore of ≥ 1 or an absolute subscore ≤ 1. Data were imported into R and analyzed using limma (v3.58.1)[6].

#### Machine learning analysis

Supervised machine learning analyses were performed to compare the predictive performance of transcriptomic and proteomic data across different tissues. Models were developed using repeated stratified 5-fold cross-validation (100 repeats), with TPOT (v0.12.2) [9] performing automated machine learning within each training fold. Feature selection was performed exclusively on the training data by ranking variables with a univariate ANOVA F-test. Five model families were evaluated: lasso, ridge, elastic-net, Gaussian Naïve Bayes, and XGBoost (v3.0.5). Performance was assessed by pooling predictions within each repeat to calculate area under the receiver operating characteristic (AUROC), generating 100 AUROC estimates per configuration. The best-performing model and feature-set size for each tissue–modality combination was selected for downstream comparison. AUROC with 95% bootstrap confidence intervals were reported.

### REFERENCES

1	Andrews S. FastQC: A Quality Control Tool for High Throughput Sequence Data. 2010.

2	Dobin A, Davis CA, Schlesinger F, Drenkow J, Zaleski C, Jha S*, et al.* STAR: ultrafast universal RNA-seq aligner. Bioinformatics 2013;**29**:15–21.

3	Liao Y, Smyth GK, Shi W. featureCounts: an efficient general purpose program for assigning sequence reads to genomic features. Bioinformatics 2014;**30**:923–30.

4	Love MI, Huber W, Anders S. Moderated estimation of fold change and dispersion for RNA-seq data with DESeq2. Genome Biol 2014;**15**:550.

5	Korotkevich G, Sukhov V, Budin N, Shpak B, Artyomov MN, Sergushichev A. Fast gene set enrichment analysis. bioRxiv 2021:060012.

6	Ritchie ME, Phipson B, Wu D, Hu Y, Law CW, Shi W*, et al.* limma powers differential expression analyses for RNA-sequencing and microarray studies. Nucleic Acids Res 2015;**43**:e47.

7	Melón-Ardanaz E, Veny M, Corraliza AM, Gudiño V, Garrido-Trigo A, Sanzo-Machuca Á*, et al.* Differential effects of tofacitinib on macrophage activation contribute to lack of response in ulcerative colitis patients. J Crohns Colitis 2025;**19**.

8	Toedter G, Li K, Marano C, Ma K, Sague S, Huang CC*, et al.* Gene expression profiling and response signatures associated with differential responses to infliximab treatment in ulcerative colitis. Am J Gastroenterol 2011;**106**:1272–80.

9	Olson RS. TPOT: A Tree-based Pipeline Optimization Tool for Automating Machine Learning. In: Moore JH, ed., 2016.
