## Supplementary figures and images for "Longitudinal multi-omics profiling of tofacitinib response identifies a pre-existing Janus kinase-tractable inflammatory state in ulcerative colitis"

### Supplementary Figure 1

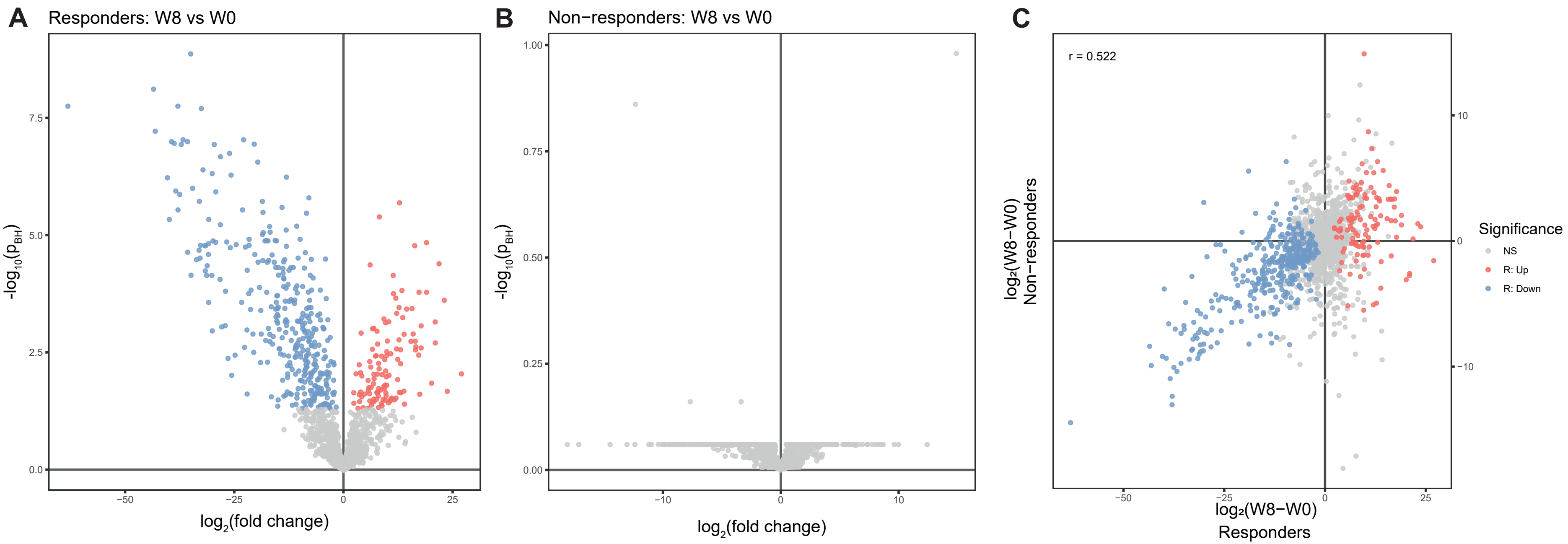

S

### Supplementary Figure 2

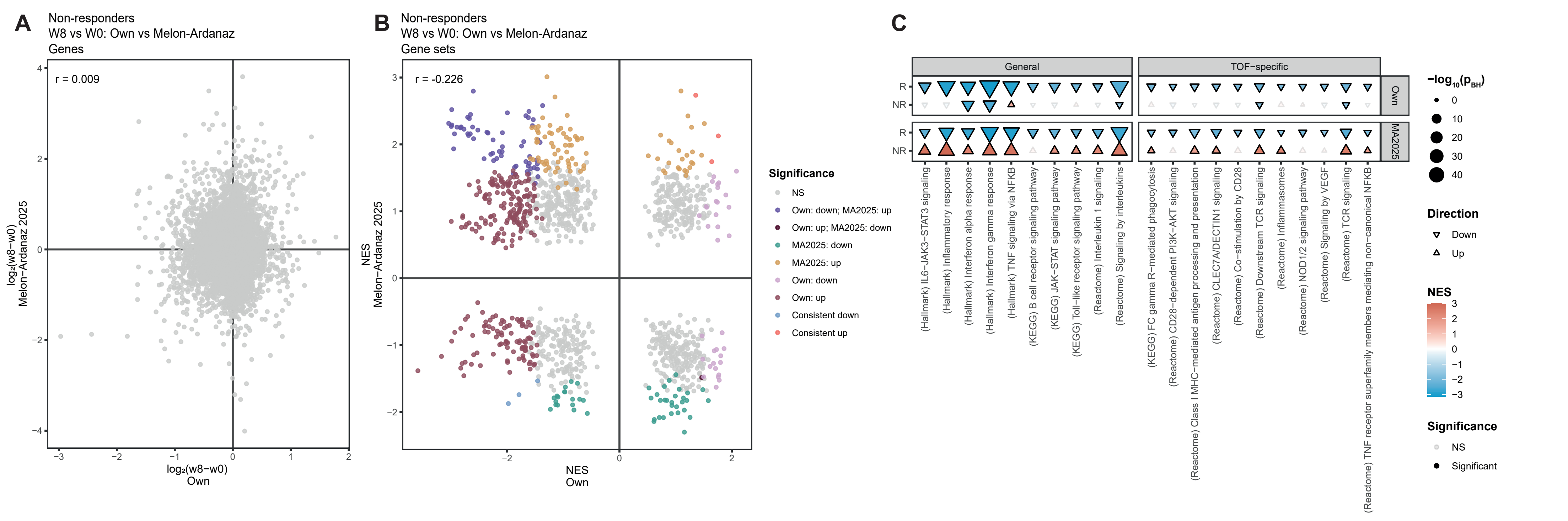

) Downstream TC
